# Longitudinal Prediction of BMI using Explainable AI: Integrating Polygenic Scores, Maternal, Early-Life and Familial Factors

**DOI:** 10.1101/2025.07.07.25331071

**Authors:** Fuling Chen, Phillip E Melton, Kevin Vinsen, Trevor Mori, Lawrence Beilin, Rae-Chi Huang

## Abstract

**Background/Objectives:** This study aimed to predict body mass index (BMI) trajectories from childhood to early adulthood using explainable artificial intelligence, integrating polygenic scores (PGS), maternal, early-life, and familial factors to identify key predictors of obesity risk and inform prevention strategies.

**Subjects/Methods:** We analysed longitudinal data from the Raine Study Gen2 cohort, recruiting 2 868 participants. This observational study, without randomization or case-control design, collected BMI measurements at ages 8, 10, 14, 17, 20, 23, and 27 years. We applied Kolmogorov-Arnold Networks (KAN) alongside conventional machine learning models, integrating epidemiological variables (maternal and paternal anthropometrics, parental education, early-life skinfold measurements) with seven BMI-related PGS. The analysis spanned from childhood to early adulthood, with no intervention administered.

**Results:** The KAN model, combining epidemiological and PGS data, achieved predictive performance with R² ranging from 0.81 at age 8 to 0.34 at age 27. BMI z-score at age 5 was the dominant predictor in early years, with PGS influence increasing post-adolescence. Maternal and paternal anthropometric measures, parental education, and early-life skinfold measurements were significant contributors. The interpretable KAN model revealed the dynamic interplay of genetic and environmental factors, with early-life BMI z-score and PGS emerging as key drivers of BMI trajectories across life stages.

**Conclusions:** These findings highlight the dynamic interplay of genetic and environmental factors across life stages, underscoring the potential of early-life BMI as a biomarker for obesity risk. Our interpretable model offers actionable insights for targeted obesity prevention strategies.

## 1. Introduction

Obesity, defined as an excessive accumulation of body fat that poses health risks, has reached epidemic proportions globally. According to the World Health Organization (WHO), worldwide adult obesity has more than doubled since 1990, and adolescent obesity has quadrupled ^1^. In Australia, recent data indicate that nearly two-thirds of adults and one in four children are overweight or obese, highlighting the urgent need for targeted public health interventions.

The transition from childhood to adulthood represents a critical period for obesity prevention. Childhood obesity significantly increases the likelihood of remaining obese into adulthood. Longitudinal studies indicate that approximately 55% of obese children remain obese in adolescence, 80% of obese adolescents continue to be obese as adults, and 70% maintain obesity over age 30 ^2^. These findings underscore the long-term health implications of childhood obesity and the importance of early intervention.

A systematic review has identified several key factors influencing obesity development from childhood to early adulthood ^3^. These include environmental, biological, pre- and postnatal, and psychosocial factors. A meta-analysis revealed that children with obese parents face a significantly higher risk of developing obesity ^4^, with this relationship being stronger in older children compared to younger ones. Parental weight exerts a significant influence on offspring, encompassing strong genetic and epigenetic effects, as well as shared environmental and social influences ^4^.

Economic factors also play a critical role in impacting children’s physical activity levels and determining the affordability and accessibility of healthy food options ^5^. Pre- and postnatal factors such as maternal pre-pregnancy obesity ^6^, excessive gestational weight gain, gestational diabetes ^6,7^, and maternal smoking before and during pregnancy ^8,9^ are significant contributors. Early childhood factors, including birth weight, rapid postnatal weight gain ^10,11^, and breastfeeding duration ^12^, have also been linked to obesity risk, emphasizing the importance of early-life interventions.

Body mass index (BMI) serves as a critical surrogate measure of obesity risk at the population level ^13^. Our study is motivated by the need to deepen our understanding of the longitudinal impact of a range of factors that influence BMI from childhood to adulthood, incorporating insights from early childhood, maternal, familial and genetic factors. Unlike previous studies, which often focus on isolated factors or lack comprehensive interpretability, we proposed a novel machine learning (ML) model that balances predictive performance with robust interpretability. Additionally, we introduced an innovative methodology to systematically identify and quantify the importance of influential factors, transforming these insights into precise BMI estimations.

## 2. Materials and methods

### a) Study population

This study used data from the Raine Study Gen2^1^. The Raine Study is a large, well-established longitudinal cohort designed to track health and developmental outcomes from pregnancy into adulthood ^14^.The Raine Study initially recruited 2,900 pregnant women (Generation 1, Gen1) and followed 2 868 children (Generation 2, Gen2). The Gen2 cohort has been followed up longitudinally from birth into early adulthood. making the study population representative of the regional population ^14^.

### b) Outcome measure

Gen2 underwent phenotyping for BMI at ages 8, 10, 14, 17, 20, 23, and 27. Weight was measured using standardized scales, and height was recorded using a stadiometer. BMI was calculated as weight (kg) divided by height squared (m²) and served as the primary outcome variable. Descriptive statistics for BMI across these age groups are presented in Supplementary 1.

### c) Epidemiological Predictors (dataset 1)– maternity, early childhood and family variables

Dataset 1 includes 201 raw variables covering maternal characteristics (including pregnancy health, maternal anthropometric conditions, maternal lifestyle, and socioeconomic factors), early childhood physical measures (including neonatal, birth, early childhood anthropometric measurements, and environmental exposures), and familial demographics (including paternal characteristics, household socioeconomic factors, and family structure and dynamics). The data processing involved multiple steps to enhance quality and ensure robust analyses, as shown in Supplementary 2. We excluded cases with congenital anomalies or multiple births to focus on typical developmental trajectories. Variables with more than 50% missing values were removed, and cases without target BMI data were filtered out. Key BMI-related measurements, such as weight and height at birth and ages one and five, were retained to calculate standardized BMI z-scores using CDC Extended BMI-for-age growth charts for very high BMI values at age five ^16^ and World Health Organization’s growth standards at birth and age one ^17^.

A correlation-based clustering algorithm grouped highly correlated variables, followed by importance-based variable selection to retain only the most predictive features for BMI outcomes (supplied in the Supplementary 2). The final steps involved excluding cases missing key variables and forming clusters, resulting in an optimized dataset for analysis. A detailed depiction of the data flow, sample sizes, and variable counts at each stage is provided in Supplementary 2, while Supplementary 1 includes variable descriptions and descriptive statistics.

### d) Genetic Factors (dataset 2)

Seven published BMI polygenic scores (PGS1-7) found in the literature were included ^18–24^ The details of the PGS were provided in Supplementary 1. These scores were generated using the pgsc_calc pipeline ^25^, which computes PGS by matching genetic variants from scoring files published in the Polygenic Score Catalog with those in the target dataset.

Preprocessing applied to both epidemiological factors (Dataset 1), and genetic factors (Dataset 2), with workflow as shown in Supplementary 2. The final numbers of preprocessing clusters were 15, 15, 25, 25, 14, 13 and 13 for the age groups of 8, 10, 14, 17, 20, 23, and 27 years, respectively.

### e) Machine learning models

The most commonly used machine learning models, including three ensemble models (Extreme Random Forest ^26^, Extreme Gradient Boosting ^27^, and Gradient Boosting Machines ^28^ and three linear models (Elastic Net ^29^, Lasso ^30^, and Ridge ^31^ regression), were used in this study. They were selected for their ability to capture complex relationships in data, offering strong predictive performance and interpretability ^32–34^.

Recently developed Kolmogorov-Arnold Networks (KAN), a deep learning model claims to enhance the accuracy of the model compared to the previous methods ^35^ with greater computational efficiency. KAN was applied to provide a formula of the relationship and explicitly present the weights of variables across layers, offering insights into the model’s decision-making process.

### f) Model training and evaluation metrics

All seven models were deployed across Dataset 1, Dataset 2, and their combined datasets. A 5-fold cross-validation strategy with varying randomization was implemented to split the data into training and testing sets. Each model was finely tuned and trained on the training set, evaluated on the testing set, with predictions collected from all five testing folds for further analysis. This approach ensured robust evaluation by mitigating the risk of overfitting and providing reliable performance estimates.

For formularization, we utilized the default formula library to derive formulas for calculating the final predictions. This approach assumes no prior knowledge of the input variables distributions or their relationships to the final outcomes.

Model performance was evaluated using the following key metrics: the Coefficient of Determination (R²) score, Root Mean Squared Error (RMSE), Mean Absolute Percentage Error (MAPE) to assess explained variance, and a confusion matrix to classify BMI predictions into four categories for the age groups at 17, 20, 23 and 27 years: underweight (BMI < 18.5), healthy weight (18.5 ≤ BMI < 25), overweight (25 ≤ BMI < 30), and obese (BMI ≥ 30). This dual evaluation provided both continuous and categorical insights into the models’ predictive capabilities. The best-performing model and dataset were selected for subsequent results presentation and analysis. Feature importance was assessed using the best model across all seven age groups.

The performance of the KAN models with formularization was compared to the original KAN models and seven conventional models by examining R² scores. Additionally, the estimates of the KAN models, both with and without formularization, were compared against the true BMI values.

To mathematically characterize the decision-making process of KAN models, we derived explicit formulas for their target outcome estimations. This was achieved by implementing the activation functions recommended by the KAN package without imposing prior functional form assumptions or distribution constraints. Instead, the functions were selected based on their optimal performance within the available function library. We have previously analysed key factors and their derived formulas identified by the KAN models—such as year 5 BMI z-scores (Y5BMIz) and the polygenic score of PGS7 ^20^—for their role in predicting the target outcomes ^36^. These demonstrate the justification of formularization of these relationships.

## 3. Results

### a) Models and datasets comparison

**Table 1** presents the R² values of seven models trained on Dataset 1 (epidemiological variables), Dataset 2 (genetic variables), and their combination across the seven age groups. Among these models, the KAN models consistently outperformed the others across all the age groups. When comparing the impact of datasets on BMI estimation, the combined use of Dataset 1 (epidemiological variables) and Dataset 2 (genetic variables) yielded the best results, followed by Dataset 1 alone. In contrast, using Dataset 2 in isolation provided little value in predicting BMI at later ages. Notably, combining both datasets significantly improved predictions, particularly for age groups over 17 years, agreed by all the seven models.

**Table 1.**
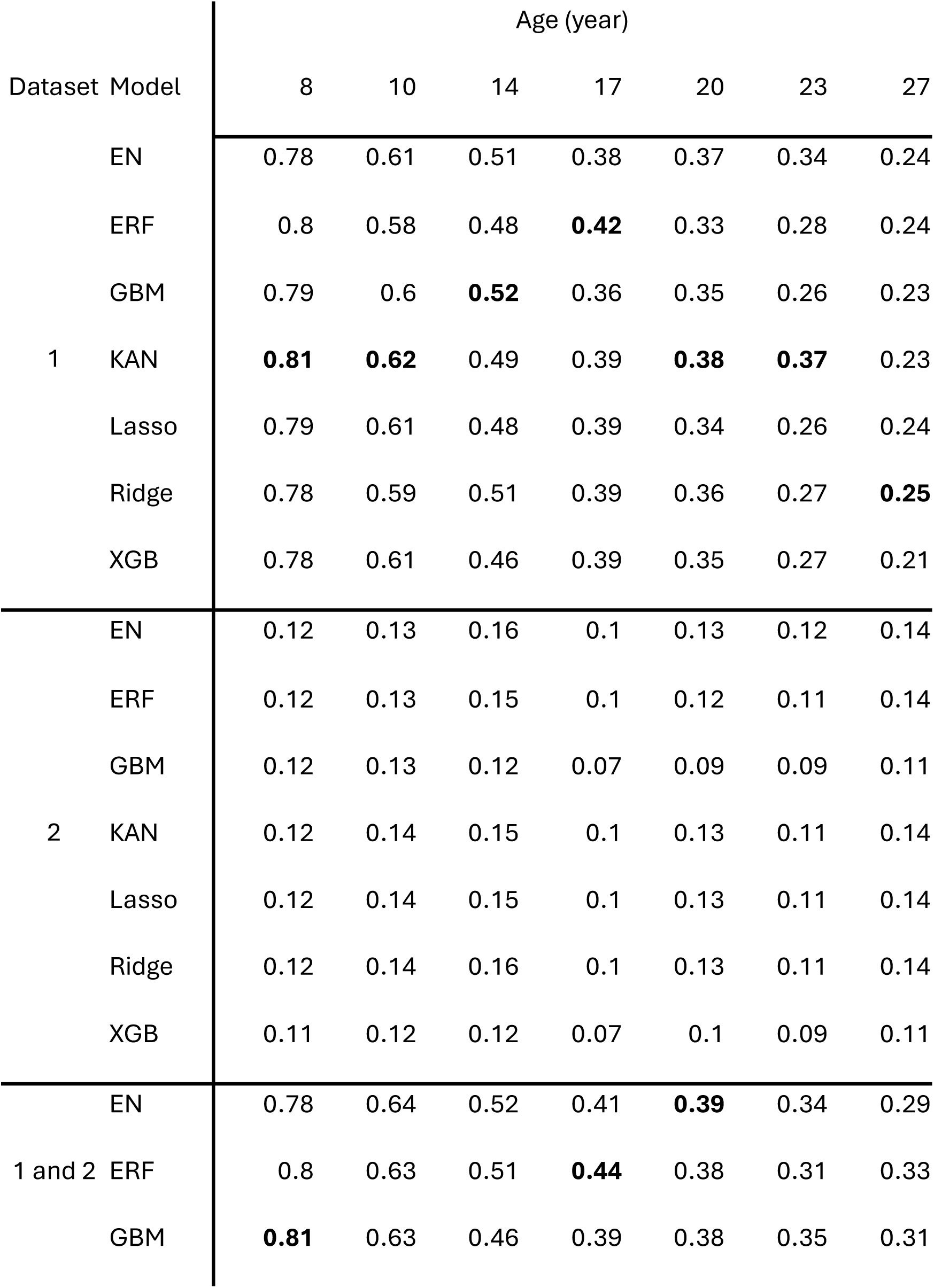
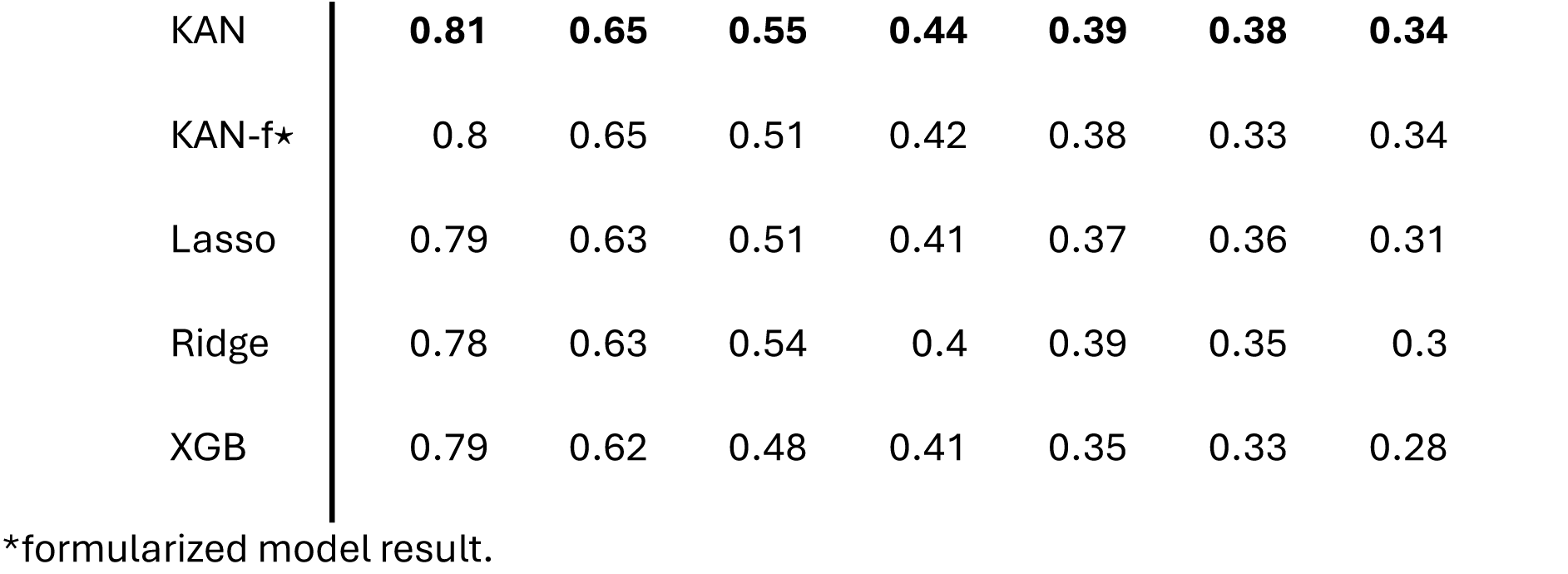
Model performance (R2) across the seven age groups and the seven models, by using Dataset 1, 2 and the combination datasets.

| Dataset Model |  | Age (year) |  |  |  |  |  |  |
| --- | --- | --- | --- | --- | --- | --- | --- | --- |
|  |  | 8 | 10 | 14 | 17 | 20 | 23 | 27 |
| 1 | EN | 0.78 | 0.61 | 0.51 | 0.38 | 0.37 | 0.34 | 0.24 |
|  | ERF | 0.8 | 0.58 | 0.48 | **0.42** | 0.33 | 0.28 | 0.24 |
|  | GBM | 0.79 | 0.6 | **0.52** | 0.36 | 0.35 | 0.26 | 0.23 |
|  | KAN | **0.81** | **0.62** | 0.49 | 0.39 | **0.38** | **0.37** | 0.23 |
|  | Lasso | 0.79 | 0.61 | 0.48 | 0.39 | 0.34 | 0.26 | 0.24 |
|  | Ridge | 0.78 | 0.59 | 0.51 | 0.39 | 0.36 | 0.27 | **0.25** |
|  | XGB | 0.78 | 0.61 | 0.46 | 0.39 | 0.35 | 0.27 | 0.21 |
| 2 | EN | 0.12 | 0.13 | 0.16 | 0.1 | 0.13 | 0.12 | 0.14 |
|  | ERF | 0.12 | 0.13 | 0.15 | 0.1 | 0.12 | 0.11 | 0.14 |
|  | GBM | 0.12 | 0.13 | 0.12 | 0.07 | 0.09 | 0.09 | 0.11 |
|  | KAN | 0.12 | 0.14 | 0.15 | 0.1 | 0.13 | 0.11 | 0.14 |
|  | Lasso | 0.12 | 0.14 | 0.15 | 0.1 | 0.13 | 0.11 | 0.14 |
|  | Ridge | 0.12 | 0.14 | 0.16 | 0.1 | 0.13 | 0.11 | 0.14 |
|  | XGB | 0.11 | 0.12 | 0.12 | 0.07 | 0.1 | 0.09 | 0.11 |
| 1 and 2 | EN | 0.78 | 0.64 | 0.52 | 0.41 | **0.39** | 0.34 | 0.29 |
|  | ERF | 0.8 | 0.63 | 0.51 | **0.44** | 0.38 | 0.31 | 0.33 |
|  | GBM | **0.81** | 0.63 | 0.46 | 0.39 | 0.38 | 0.35 | 0.31 |

|  |  |  |  |  |  |  |  |
| --- | --- | --- | --- | --- | --- | --- | --- |
| KAN | **0.81** | **0.65** | **0.55** | **0.44** | **0.39** | **0.38** | **0.34** |
| KAN-f★ | 0.8 | 0.65 | 0.51 | 0.42 | 0.38 | 0.33 | 0.34 |
| Lasso | 0.79 | 0.63 | 0.51 | 0.41 | 0.37 | 0.36 | 0.31 |
| Ridge | 0.78 | 0.63 | 0.54 | 0.4 | 0.39 | 0.35 | 0.3 |
| XGB | 0.79 | 0.62 | 0.48 | 0.41 | 0.35 | 0.33 | 0.28 |

\*formularized model result.

Regarding the influence of age, all models agreed that the performance gradually decreased with age, ranging from R^2^ of 0.81 at the age of 8 years to 0.34 at the age of 27 years. Notably, the formularized models exhibited performance that closely aligned with their original counterparts, showing minimal differences. The other metrics, including RMSE, MAPE, and the confusion matrices of the four classes are provided in Supplementary 2.

As the KAN models trained on the combined datasets yielded the best results, the subsequent results and analyses will focus on the KAN models utilizing both datasets 1 and 2.

### b) Overall feature importance

The feature importance in the decision-making process of the KAN model was determined by multiplying the weights of each activation function from the leaf nodes (input variables) to the output (the BMI values of a target age group), with the visualised pruned tree plots shown in Supplementary 2 for the seven age groups.

As shown in Figure 1, the weights exhibit significant variation across different age groups and variables. Most features contribute relatively low weights to the predictions. The most influential variable is the BMI z-score at 5 years of age. It demonstrates its dominance in early age groups but shows a marked decline in importance after age 20. While there is decline, at 27 years of age, it maintains similar absolute weight as several of the polygenic risk scores. In contrast, polygenic scores (PGS), including PGS1, PGS3, PGS5, PGS6, and PGS7, have a lower impact in early age groups but gradually catch up with Y5BMIz after age 17. This shift aligns with the findings in detailed above in Section 3.a, where the R² scores benefit more from using both datasets rather than Dataset 1 alone, starting from age 17.

**Figure 1.**
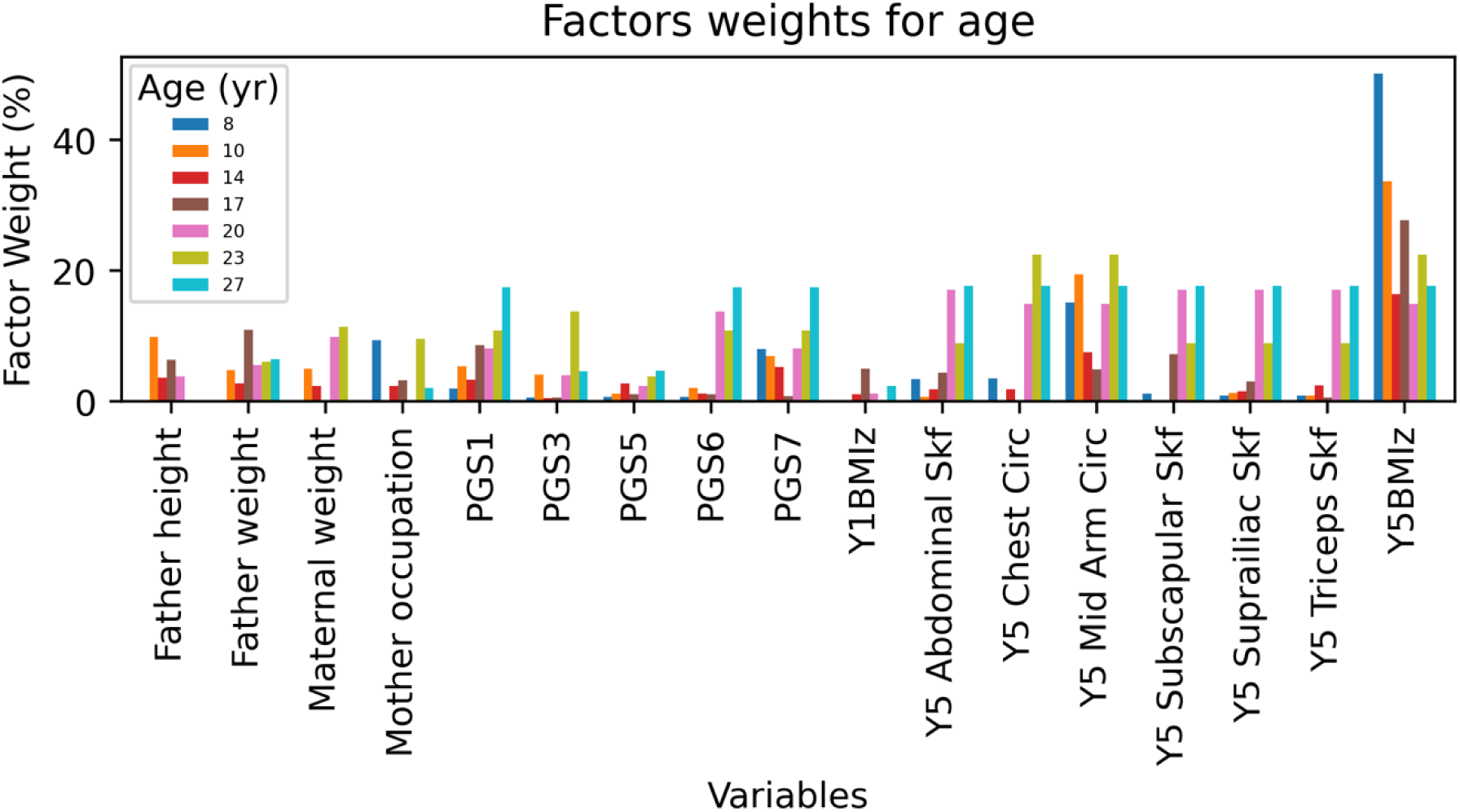
Key variables and their weights (in percentages) across seven age groups. Only variables appearing in at least four age groups are shown. The polygenic scores 1-7 represent PGS1 ^19^ PGS2 ^23^ PGS3 ^24^ PGS4 ^22^ PGS5 ^21^ PGS6 ^18^ PGS7 ^20^. PGS2 and PGS4 are not presented due to their high correlations with PGS1 and PGS3.

Other anthropometric measurements, including abdominal, suprailiac, subscapular and triceps skinfold measurements and arm and chest circumference at age 5, follow a similar trend in the models, with their weights increasing across the age groups. Variables consistently predicting offspring BMI across all age groups include maternal occupation, pre-pregnancy weight, late-pregnancy weight, maternal height-to-weight ratio, paternal weight, and paternal height. Additional variables, selected as important for specific ages not shown in Figure 1, include pre-pregnancy and pregnancy smoking (ages 8, 10, 14), paternal occupation (ages 10, 14, 27), maternal age (ages 14, 17, 27), childcare (ages 14, 17, 23), delivery mode (age 14), placenta weight (ages 14), birth BMI z-score (age 17), solid food introduction age (age 17), paternal education (age 20), sex (age 20), breastfeeding (age 23), birth anthropometrics (age 23), Apgar score (age 23), maternal height (age 23), paternal age (age 27), and birth month (age 27). Further details are provided in Supplementary 1.

### c) Formularization

Our results demonstrate that the models maintain comparable performance after formula derivation, as shown in Table 1 (“KAN-f*”). This consistency enables further exploration of the mathematical relationships between key variables and their role in predicting BMI. The derived formulas for BMI estimation as a function of the selected variables across the seven age groups are provided in Supplementary 1.

To identify and analyse the most influential factors across age groups, we examined the formula representations of Y5BMIz and the most heavily weighted polygenic score PGS7 ^20^ along with their relationships with actual BMI values.

The BMI estimation and the formula representation of BMI z-score at 5 years old (Y5BMIz) across the seven age groups are demonstrated in Supplementary 2. Notably, Y5BMIz is one of 10 to 20 key factors utilized by the KAN model in its final decision-making process. Before age 20, the derived function of Y5BMIz follows an exponential or first-quadrant sine function, a pattern reflected in both actual BMI values and model estimations. After age 20, however, this relationship weakens, and the model struggles to formulate a stable mathematical representation for Y5BMIz. Observations of actual BMI values beyond age 20 reveal a high concentration of data points in the lower range of both the Y5BMIz factor and the true BMI, indicating data sparsity. This sparsity limits the KAN model’s ability to derive a robust formula and estimation for Y5BMIz. Nevertheless, since the model incorporates multiple factors during training, it leverages the additional variables to maintain predictive performance.

Beyond Y5BMIz, we observed that the influence of polygenic scores (PGS) on BMI increased with age. Using PGS7 as a representative example, we identified a consistent positive correlation between PGS7 and BMI across all seven age groups. To mathematically characterize this relationship, we applied the KAN model to derive functional representations of PGS7. The figures in Supplementary 2 present BMI estimations alongside the formulaic representations of PGS7 across the seven age groups. Most derived functions effectively capture the positive correlation between PGS7 and BMI, except at age 17, where PGS7 carries a lower weight.

## 4. Discussion

This study has explored greater than 200 variables that are relatively accessible and often routinely collected from demographic, family history, antenatal history, early life and increasingly available genetic SNP array data, to predict those at risk of future overweight and obesity. Using KAN machine learning methods, formulas of the algorithm to calculate the risk are produced alleviating the “black box” issue of previous machine learning methods, problematic in using machine learning in spheres where clinical decisions are being made. We find that in predicting BMI in younger age groups, factors such as maternal weight during pregnancy, paternal height, and anthropometric measures at age 5 years, exhibit strong associations with BMI. As individuals transition through adolescence into early adulthood, the influence of these early-life factors diminishes, while genetic factors, captured through PGS, become increasingly significant.

These findings align with existing literature highlighting the critical role of early-life conditions in shaping long-term health outcomes ^37^. Factors such as maternal weight during pregnancy, paternal height, paternal weight, and early-life anthropometric measures exhibit strong associations with BMI across the ages examined in this study.

As individuals transition from adolescence into early adulthood, the influence of these early-life factors diminishes, while genetic factors, represented by PGS, become increasingly significant. Although PGS alone may not be sufficient for accurate BMI prediction, its predictive power is greatly enhanced when integrated with early-life and environmental data. This synergistic effect becomes particularly evident from age 17 onward, where combining genetic and early-life variables significantly improves model performance.

These findings underscore the dynamic interplay between genetic predisposition and environmental influences in shaping BMI across different life stages. Given that the PGS used in this study were derived from external data cohorts and have been independently validated, our results strongly support integrating genetic data with epidemiological variables in longitudinal studies to better characterize the factors influencing BMI.

Anthropometry at age 5 years old consistently emerges as the most influential predictor across all studied age groups. Its exponential relationship with BMI in later life underscores its potential as a clinical biomarker for assessing the risk of overweight and obesity. The predictive power of Y5BMIz is particularly pronounced in younger age groups, where it serves as a critical determinant of later BMI outcomes. Interestingly, at older ages, skin folds at 5 years old becomes a more powerful predictor, rather than BMI z-score at 5 years. These subtle nuances in measures of body composition provide some insights into the value of consideration of body composition, rather than BMI per se which has been debated ^38^. The majority of studies have concluded that BMI is at least as accurate as skin fold thicknesses in predicting future cardiovascular risk ^38–40^.

While, both BMI and skin fold measures are indirect measurements of adiposity, skinfold measurements are a closer measure of subcutaneous fat which in children has been shown to track in childhood ^41^. It could be hypothesized that preferential deposits of fat will results in earlier saturation of subcutaneous adipose tissue and expansion and accumulation of ectopic fat associated with metabolic dysfunction ^42^ which may not become apparent until mid-twenties and beyond. At a histopathological level, subcutaneous tissue in overweight children had greater adipocyte surface area and collagen content in their subcutaneous tissue compared to normal weight children ^43^.

Even during adolescence and early adulthood, year 5 adiposity continues to exert a significant influence. This persistent relevance suggests that early-life BMI not only reflects immediate health conditions but also captures latent risk factors that manifest later in life. Consequently, BMI z-score at 5 years of age could serve as a valuable tool in early intervention strategies, enabling healthcare professionals to identify individuals at risk of developing obesity and implement targeted preventive measures. The concept of critical time points in development for future risk of disease has been expounded with a lifecourse lens on developmental origins of health and disease ^44^. Critical time points have been included the first 1000 days and adolescence. Our current data suggest that BMI as early as approximately 5 years of age is a strong predictor of future obesity. This is consistent with other studies of longitudinal growth ^45^ which show that from approximately 5 years old onwards, tracking of obesity is stable. Prior to that, there are some groups of individuals who are on obesity trajectories who proceed from being relatively undersized at birth (presumably with some in utero insult such as malnutrition or smoking during pregnancy) who develop “catch up growth” and ultimate high risk for obesity ^46^.

There are some limitations from our study design in making such categorical conclusions. These include that specific time points of anthropometry collection which do not allow for assessment of data collected at time points other than the stipulated birth, 2, 3 and 5 years. However, we see that despite many variables included from these time points, none were used in the final machine learning models. We also temper this conclusion stating that it refers to prediction on a population scale attempting to identify a cross-sectional age early life that reliably predicts future obesity. Naturally risk of an individual may be better predicted with velocity of BMI change with repeated measurements.

In this study, we chose to develop unified models for BMI prediction across both sexes. While sex was included among the initial variables, feature selection identified it as a significant predictor only at age 20, where it exhibited a relatively lower weight compared to dominant factors such as early-life BMI z-scores and polygenic scores. Across other age groups, sex did not consistently emerge as a critical determinant of BMI trajectories, indicating that its influence is secondary to other predictors. Secondly, constructing separate sex-specific models would reduce the sample size for each analysis, potentially limiting the statistical power and generalizability of the findings. Lastly, early childhood BMI z-scores, calculated using sex-specific standards, already incorporate sex-related differences in growth patterns, ensuring that these variations are adequately accounted for in our models. By prioritizing a unified approach, we maximized the robustness of our predictions and focused on the most influential factors driving BMI across the life course.

Parental factors also play a pivotal role in shaping BMI outcomes. Our findings highlight the contributions of variables such as parental education, parental height and weight, maternal age at childbirth, and maternal weight gain during pregnancy. These factors, which likely encapsulate both genetic and environmental influences, exhibit consistent associations with BMI across all studied age groups.

The education level of parents, for instance, reflects broader socioeconomic and environmental conditions that influence health behaviours and access to resources. Similarly, maternal weight gain during pregnancy may indicate nutritional and metabolic conditions that impact fetal development. The observed associations between these factors and BMI underscore the importance of considering a holistic framework that integrates genetic, maternal, and environmental influences in BMI prediction models.

Notably, the interplay between these factors evolves with age. For example, while maternal and paternal anthropometric measures show limited influence during early childhood, their impact increases in adolescence and early adulthood. This shift aligns with the growing importance of genetic factors, as reflected in the increasing contribution of PGS variables during these later stages.

This study is a comprehensive attempt to integrate using new machine learning methodology prospectively collected genetic and epidemiological data with longitudinal sample collection of over close to three decades.

Limitations of this study are that data are collected at fixed time points according to original study design, hence no extrapolation to different ages can be made. A further point of qualification is there is no intent to imply causation. Indeed, certain variables may have greater level of direct causation but not be selected whereas others with indirect causation selected because they encapsulate greater latent variance. Moreover, the use of BMI as the sole indicator of obesity risk presents a limitation. Although BMI is a widely accepted and consistently applied measure in obesity research, it may not fully capture the complexity of adiposity. Alternative anthropometric measures—such as waist circumference, waist-to-hip ratio, or waist-to-height ratio—could provide complementary insights into obesity-related health risks. However, BMI was chosen as the primary indicator due to its established reliability, availability within the dataset, and alignment with population-level standards for assessing obesity risk.

Future studies should follow up these findings to ascertain if genetics become increasingly influential at predicting BMI at older ages and whether skinfold thickness (subcutaneous fat thickness measurements) at 5 years maintains or even increases in importance. The findings underscore the importance of age, early-life factors, and parental and maternal influences in shaping BMI trajectories from childhood to early adulthood. These insights pave the way for more targeted and effective interventions aimed at promoting healthy weight and preventing obesity over the life course.

## Acknowledgements

We gratefully acknowledge all Raine Study participants and their families for their continued participation in the study, as well as the Raine Study team for study co-ordination and data collection. We also thank the NHMRC and the Raine Medical Research Foundation for their support. The core management of the Raine Study is funded by The University of Western Australia, Curtin University, The Kids Research Institute Australia, Women and Infants Research Foundation, Edith Cowan University, Murdoch University, The University of Notre Dame Australia and the Western Australian Future Health Research and Innovation Fund (2023-2024; Grant ID WACSOSP2023-2024). The Pawsey Supercomputing Centre provided computation resources to carry out analyses required with funding from the Australian Government and the Government of Western Australia. The data collection of the Raine Study Gen1- and 2-1, 2, 5, 8, 10, 14, 17, 20, 22, and 26 year follow-ups were funded by NHMRC project grants (211912, 003209, 572613, 403981, 353514, 572613, 403981, 1059711, 1084947), and The Raine Medical Research Foundation.

## Author Contributions

F.C. designed and conducted the study, developed the model, aggregated and visualized the results, performed the analysis, and drafted the manuscript and supplementary materials. R.H. was responsible for data acquisition, results interpretation, analysis, and manuscript writing. P.M. contributed to data acquisition, model evaluation, and results interpretation and analysis. All authors reviewed and approved the final manuscript.

## Competing Interests

The authors declare no competing financial interests.

## Data Availability

The datasets generated during and/or analysed during the current study are not available. The Raine study is committed to a high level of confidentiality of the data in line with the informed consent provided by participants. Requests for data should be directed to the Raine Study Executive.

## Footnotes

1 The variables in the Rain Study are mostly harmonized with the LifeCycle Project-EU Child Cohort Network ^15^

## Notes

### Competing Interest Statement

The authors have declared no competing interest.

### Funding Statement

This study did not receive any funding.

### Author Declarations

Dr Fuling Chen The University of Western Australia Dear Dr Chen HUMAN RESEARCH ETHICS EXEMPTION FROM REVIEW. Early Life Origins of Cardiovascular Health: A Machine Learning-Driven Risk Prediction Algorithm for Pediatric Populations Based on the information you have provided to the Human Ethics office in relation to the above project, the described activity has been assessed as exempt from ethics review at the University of Western Australia. However, should there be any significant changes to the project, you must contact the HREO to determine whether your exempt status remains valid or whether you will be required to submit an application for ethics approval. If you have any queries please contact the Human Ethics office at. Please ensure that you quote the file reference 2025/ET000396 and the associated project title in all future correspondence. Yours sincerely Shihab Mundekkadan Senior Officer Human Ethics and Clinical Trials

