## Supplementary 2 for "Longitudinal Prediction of BMI using Explainable AI: Integrating Polygenic Scores, Maternal, Early-Life and Familial Factors"

### Supplementary 1. Data Processing

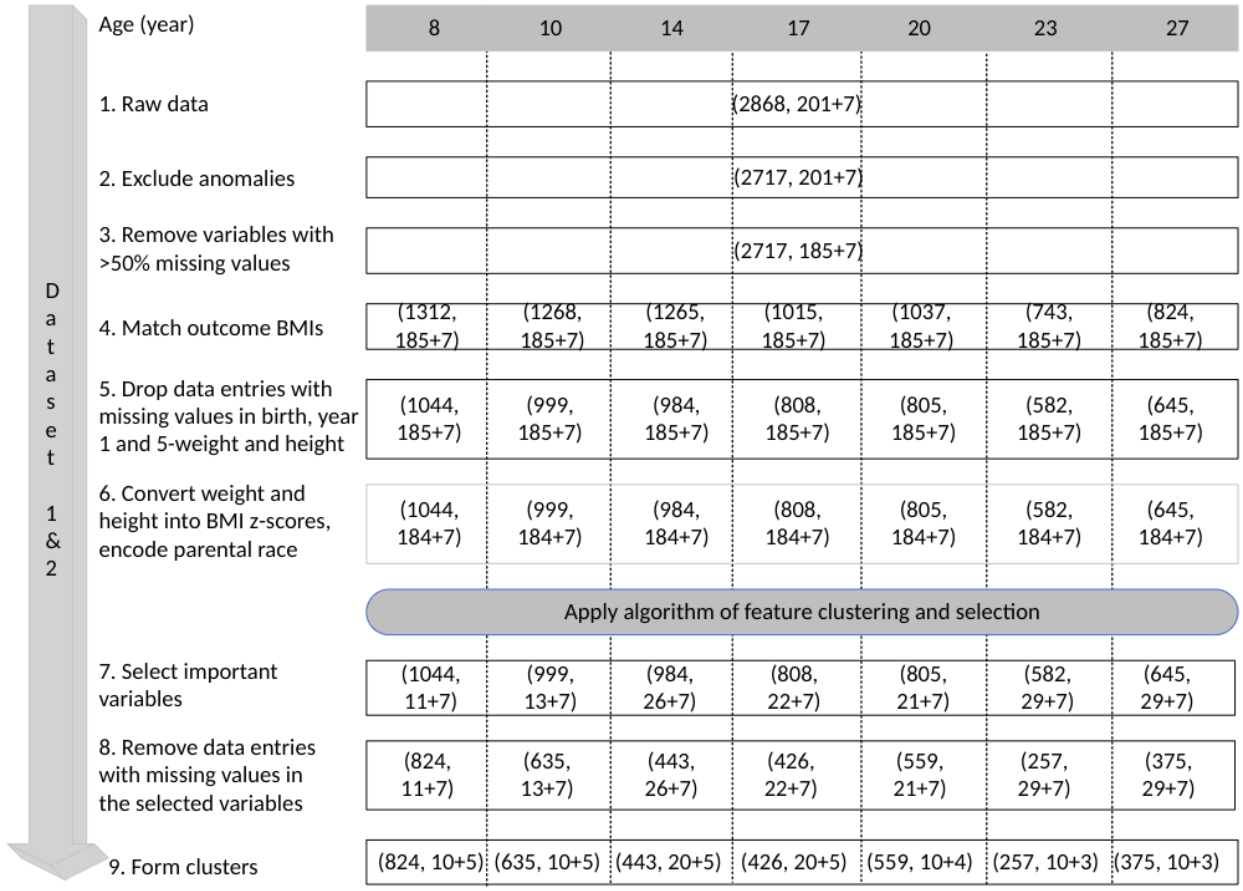


Supplementary Figure 1. Datasets preprocessing workflow. The form of (i, j+k) represents the number of samples (i), the number of dataset 1 variables (j), and the number of polygenic scores of dataset 2 (k).

### Supplementary 2. Algorithm of variables clustering and selection

To optimize data cleansing while preserving as many entries as possible, we developed a variable clustering and selection algorithm, applied after Step 6: *"Convert weight and height into BMI z-scores, encode parental race"* (as shown in Figure 1). This algorithm consists of three main stages: Step A (*"Deep Clean"*), Step B (*"Cluster Variables"*), and Step C (*"Select Important Clusters"*). Detailed descriptions of these steps are provided in Figure 1. The input data for this algorithm corresponds to the dataset obtained from Step 6 in Figure 1.

| Age (year) | 8 | 10 | 14 | 17 | 20 | 23 | 27 |
|---|---|---|---|---|---|---|---|
| Input data | (1044, 184+7) | (999, 184+7) | (984, 184+7) | (808, 184+7) | (805, 184+7) | (582, 184+7) | (645, 184+7) |
| DEEP CLEAN: A1. Exclude variables with >20% missing values | (1044, 181+7) | (999, 181+7) | (984, 180+7) | (808, 181+7) | (805, 181+7) | (582, 181+7) | (645, 181+7) |
| A2. Impute the categorical variables with >80% similarity | (1044, 181+7) | (999, 181+7) | (984, 180+7) | (808, 181+7) | (805, 181+7) | (582, 181+7) | (645, 181+7) |
| A3. Drop data entries with missing values | (176, 181+7) | (167, 181+7) | (166, 180+7) | (147, 181+7) | (148, 181+7) | (113, 181+7) | (120, 181+7) |
| A4. Remove constant variables | (176, 140+7) | (167, 140+7) | (166, 138+7) | (147, 140+7) | (148, 139+7) | (113, 139+7) | (120, 140+7) |
| B. Cluster varaibles | (171, 121) | (167, 120) | (166, 101) | (147, 120) | (148, 71) | (113, 71) | (120, 72) |
| C. Select: n_cluster | 10+5 | 10+5 | 20+5 | 20+5 | 10+4 | 10+3 | 10+3 |
| n_variable | 11+7 | 14+7 | 26+7 | 20+7 | 21+7 | 29+7 | 24+7 |
| r_cutoff | 0.9 | 0.9 | 0.8 | 0.9 | 0.6 | 0.6 | 0.6 |

Supplementary Figure 2. Workflow of variables clustering and selection of Dataset 1 and 2. The form of (i, j+k) represents the number of samples (i), the number of Dataset 1 variables (j) and the number of Dataset 2 PGS (k)

In Step A (*"Deep Clean"*), we refined the dataset by removing variables with more than 20% missing values, imputing categorical variables with over 80% similarity, and dropping data entries with missing values in the remaining variables. Additionally, constant variables were eliminated. As shown in Figure A.I.1, this process significantly reduced the number of data entries. While such a reduction could adversely impact machine learning model development if used directly for training, we utilized this "clean" data solely for clustering and selection to identify important factors and eliminate redundant variables.

The implementation of Step B (*"Cluster Variables"*) is illustrated in Figure 2. First, we computed the correlation matrix of the input variables. For any pair of items with a correlation coefficient (r(A, B)) exceeding the threshold value (*r_cutoff*), we replaced them with a single component (*X*) derived through Principal Component Analysis (PCA). Each item could represent either a single variable or a cluster of multiple variables. The *r_cutoff* value was tuned during model training and selected from options of 0.6, 0.7, 0.8, and 0.9.

After clustering, we obtained both the cluster patterns and the clustered dataset for Step C (*"Select n_cluster, n_variable, r_cutoff"*) and subsequent analyses. These clusters were treated as independent units, exhibiting minimal dependency on one another.

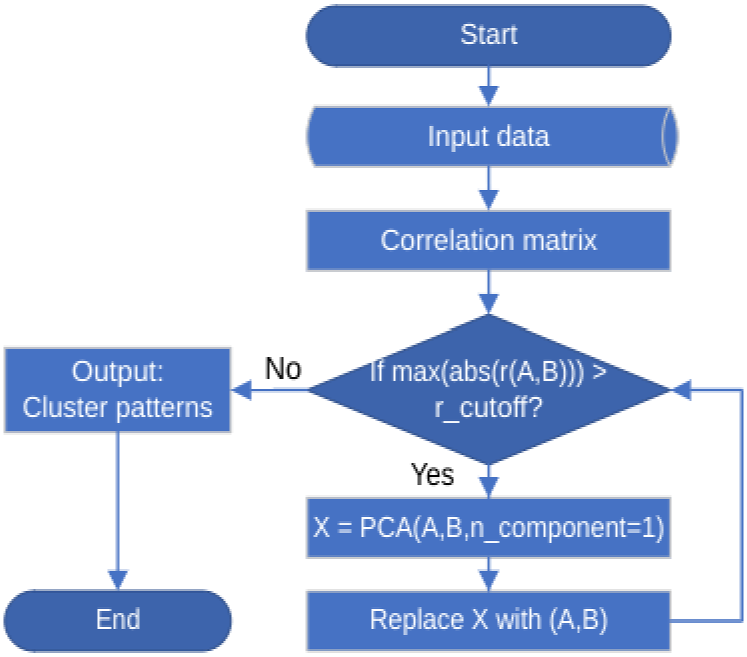

Supplementary Figure 3. Correlated-based clustering algorithm.

In Step C (*"Selection"*), we employed Recursive Feature Elimination (RFE) integrated with Extreme Random Forest (ERF) to identify potential predictors. The optimal number of predictors was determined by tuning among 10, 20, 30, and 40.

### Supplementary 3. Model performance

3.1 Root Mean Squared Error (RMSE)

RMSE were calculated in the following formula:

$$RMSE = \sqrt{\frac{1}{n}\sum_{i=1}^{n}(y_i - \overline{y_i})^2},$$

where $y_i$represents the actual values, and $\overline{y_i}$ represents the predicted values.

| Dataset | Model | Age (year) | | | | | | |
|---|---|---|---|---|---|---|---|---|
| | | 8 | 10 | 14 | 17 | 20 | 23 | 27 |
| 1 | EN | 1.2 | 2.14 | 2.84 | 2.78 | 3.7 | 3.8 | 4.64 |
| | ERF | 1.13 | 2.06 | 3 | 3.02 | 4.13 | 3.39 | 4.62 |
| | GBM | 1.14 | 2.1 | 2.76 | 3.12 | 3.72 | 3.55 | 4.83 |
| | KAN | 1.11 | 2.09 | 2.92 | 3.41 | 3.71 | 3.73 | 4.78 |
| | Lasso | 1.15 | 2.15 | 2.85 | 3.06 | 3.76 | 3.56 | 4.8 |

| | | | | | | | | |
|---|---|---|---|---|---|---|---|---|
| | Ridge | 1.17 | 2.17 | 2.85 | 3.07 | 3.67 | 3.59 | 4.62 |
| | XGB | 1.17 | 2.15 | 2.97 | 3.1 | 3.82 | 4.01 | 4.73 |
| 2 | EN | 2.38 | 3.17 | 3.85 | 4.12 | 4.70 | 4.87 | 5.33 |
| | ERF | 2.38 | 3.17 | 3.87 | 4.12 | 4.72 | 4.89 | 5.32 |
| | GBM | 2.39 | 3.18 | 3.94 | 4.18 | 4.80 | 4.94 | 5.41 |
| | KAN | 2.39 | 3.17 | 3.86 | 4.12 | 4.70 | 4.90 | 5.31 |
| | Lasso | 2.38 | 3.17 | 3.85 | 4.12 | 4.70 | 4.87 | 5.33 |
| | Ridge | 2.38 | 3.17 | 3.85 | 4.11 | 4.69 | 4.87 | 5.32 |
| | XGB | 2.39 | 3.19 | 3.92 | 4.20 | 4.78 | 4.93 | 5.40 |
| 1 and 2 | EN | 1.19 | 1.99 | 2.91 | 3.05 | 3.56 | 3.43 | 4.63 |
| | ERF | 1.09 | 2.09 | 2.65 | 2.99 | 3.59 | 3.99 | 3.76 |
| | GBM | 1.07 | 2.08 | 2.64 | 3.12 | 3.66 | 3.39 | 4.59 |
| | KAN | 1.1 | 2.01 | 2.7 | 3 | 3.96 | 3.7 | 4.46 |
| | KAN-f⋆ | 1.1 | 2.02 | 2.59 | 2.99 | 3.96 | 3.45 | 4.46 |
| | Lasso | 1.17 | 2.07 | 2.81 | 2.73 | 4 | 3.52 | 4.59 |
| | Ridge | 1.19 | 2.07 | 2.73 | 3.11 | 3.62 | 3.66 | 4.44 |
| | XGB | 1.13 | 2.11 | 2.93 | 3.08 | 3.7 | 3.42 | 4.69 |

*formularized model result.

Supplementary Table 1. Model performance (RMSE) across the seven age groups and the seven models, by using Dataset 1, 2 and the combination datasets.

3.2 Mean Absolute Percentage Error (MAPE)

RMSE were calculated in the following formula:

$$MAPE = \frac{1}{n}\sum_{i=1}^{n}\left|\frac{y_i - \overline{y_i}}{y_i}\right| \times 100\,,$$

| Dataset | Model | Age (year) | | | | | | |
|---|---|---|---|---|---|---|---|---|
| | | 8 | 10 | 14 | 17 | 20 | 23 | 27 |
| 1 | EN | 4.96 | 8.36 | 9.48 | 8.89 | 11.18 | 11.17 | 12.79 |
| | ERF | 4.64 | 8.13 | 9.81 | 9.62 | 11.89 | 9.84 | 12.08 |
| | GBM | 4.71 | 8.24 | 9.2 | 9.84 | 11.01 | 10.68 | 12.84 |
| | KAN | 4.57 | 8.36 | 9.77 | 10.41 | 11 | 10.98 | 12.61 |
| | Lasso | 4.8 | 8.48 | 9.53 | 9.67 | 11.38 | 10.66 | 12.66 |
| | Ridge | 4.98 | 8.6 | 9.55 | 9.83 | 11.17 | 10.89 | 12.68 |
| | XGB | 4.75 | 8.3 | 9.89 | 9.81 | 11.42 | 11.48 | 12.92 |
| 2 | EN | 9.76 | 12.57 | 12.96 | 12.53 | 13.44 | 13.93 | 14.27 |
| | ERF | 9.74 | 12.5 | 12.98 | 12.54 | 13.49 | 14.03 | 14.18 |

| | | | | | | | | |
|---|---|---|---|---|---|---|---|---|
| | GBM | 9.75 | 12.52 | 13.24 | 12.78 | 13.75 | 14.15 | 14.41 |
| | KAN | 9.75 | 12.43 | 12.88 | 12.55 | 13.51 | 13.89 | 14.29 |
| | Lasso | 9.76 | 12.54 | 12.97 | 12.51 | 13.43 | 13.95 | 14.29 |
| | Ridge | 9.76 | 12.53 | 12.96 | 12.5 | 13.41 | 13.94 | 14.27 |
| | XGB | 9.72 | 12.5 | 13.14 | 12.71 | 13.69 | 14.12 | 14.35 |
| 1 and 2 | EN | 4.92 | 7.92 | 9.6 | 9.58 | 10.7 | 10.31 | 12.3 |
| | ERF | 4.57 | 8.29 | 8.99 | 9.54 | 10.53 | 10.91 | 10.35 |
| | GBM | 4.53 | 8.09 | 9.07 | 9.66 | 10.72 | 10.05 | 12.03 |
| | KAN | 4.52 | 8.12 | 9.17 | 9.53 | 11.24 | 10.58 | 11.79 |
| | KAN-f⋆ | 4.59 | 8.16 | 8.98 | 9.51 | 11.24 | 10.36 | 11.79 |
| | Lasso | 4.93 | 8.3 | 9.5 | 8.6 | 11.29 | 10.27 | 12.01 |
| | Ridge | 4.95 | 8.25 | 9.18 | 9.76 | 10.7 | 10.67 | 11.91 |
| | XGB | 4.72 | 8.28 | 9.84 | 9.71 | 10.93 | 10.32 | 12.08 |

*formularized model result.

Supplementary Table 2. Model performance (MAPE) across the seven age groups and the seven models, by using Dataset 1, 2 and the combination datasets.

3.3 Confusion matrix for age 17, 20, 23 and 27 years

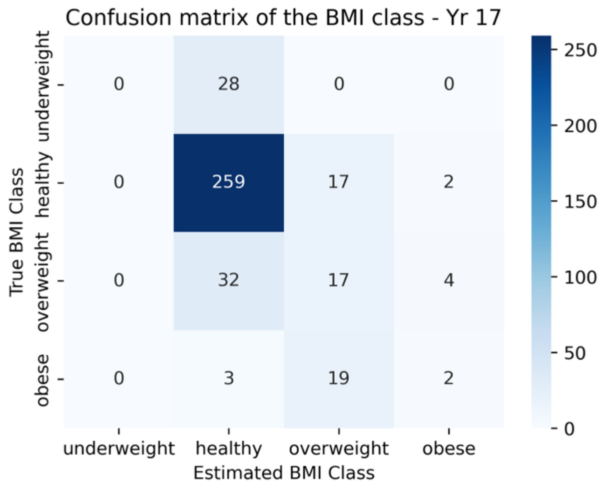


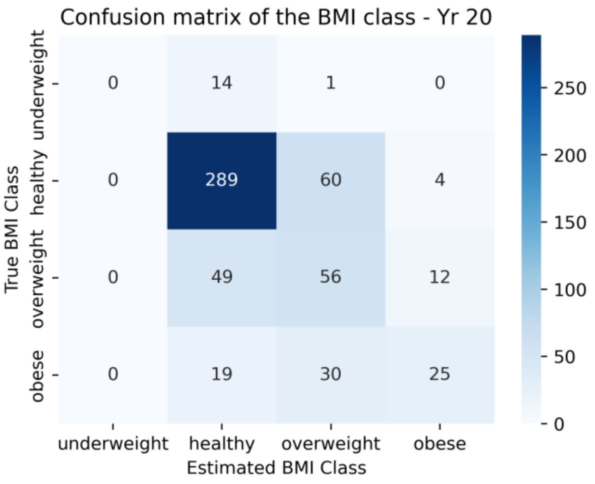

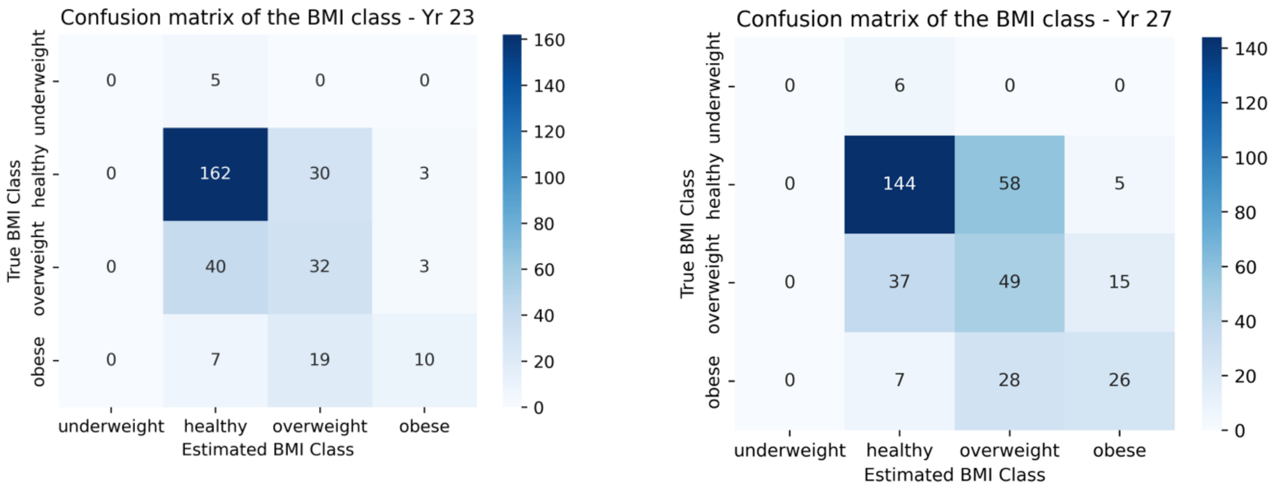


Supplementary Figure 4. Confusion matrix for age 17, 20, 23 and 27 years

### Supplementary 4. Pruned tree plots of the KAN models

The bottom black points represent the selected factors used in model development, the middle layer points represent the first-layer nodes of the KAN models, and the top point corresponds to the target BMI value for the specific age. The colour intensity of the connecting lines reflects the impact of the lower node on the upper node. Additionally, nail plots in the middle of the connecting lines represent the activation functions used in the models, with the most significant ones highlighted for further discussion in subsequent sections.

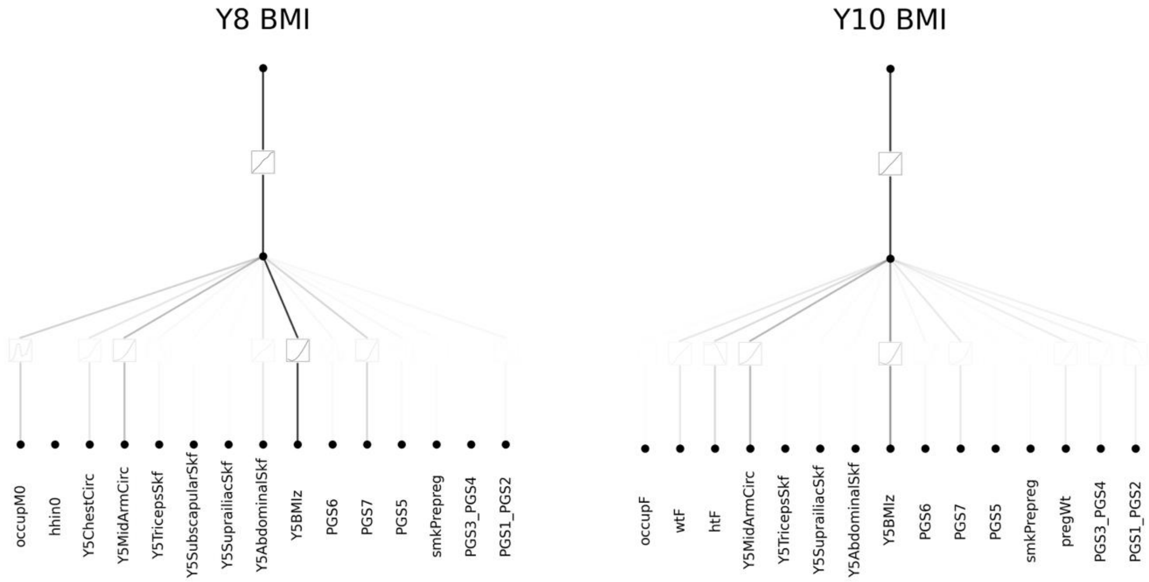

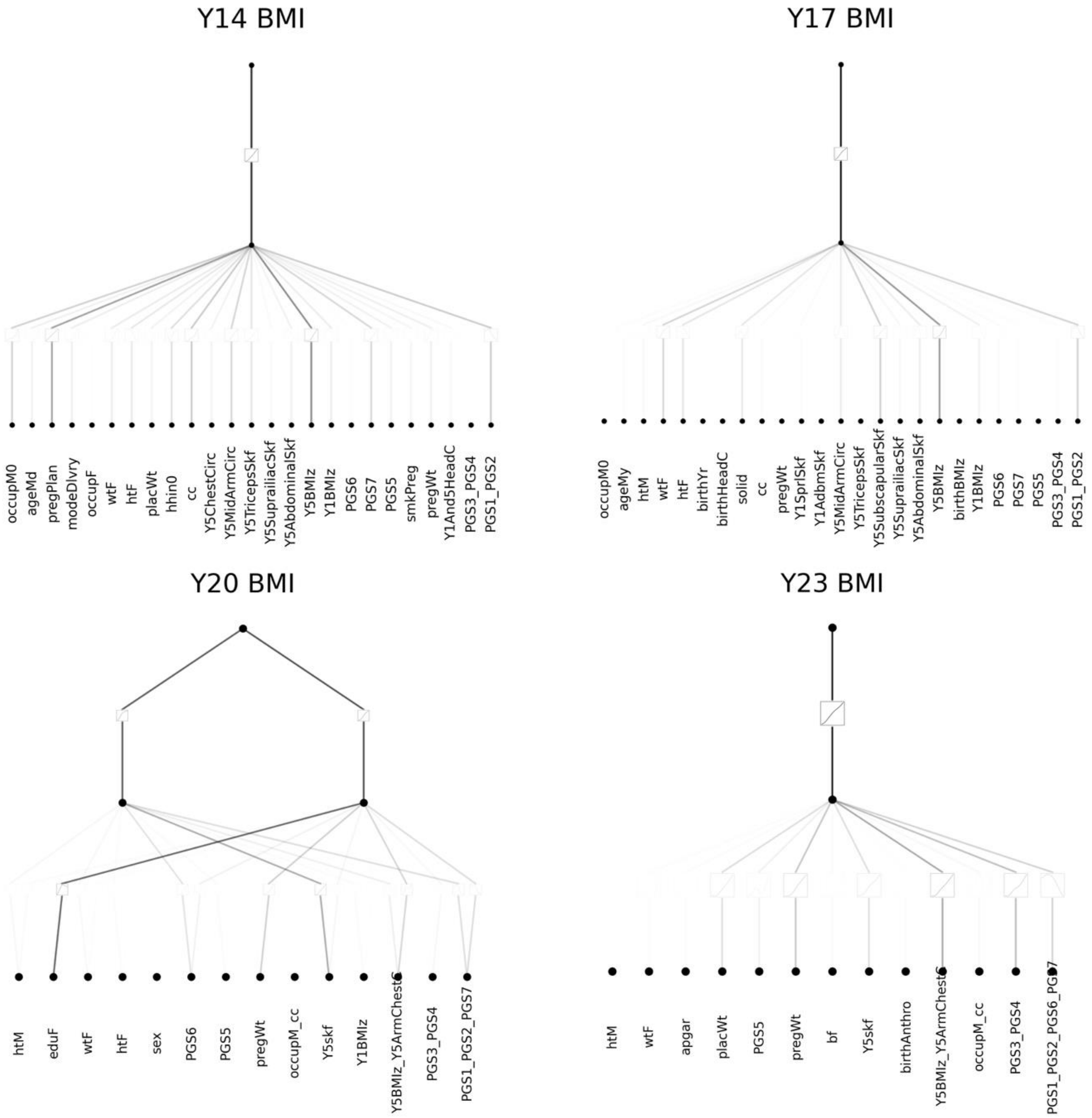
Y14 BMI
occupM0
ageMd
pregPlan
modeDlvry
occupF
wtF
htF
placWt
hhin0
cc
Y5ChestCirc
Y5MidArmCirc
Y5TricepsSkf
Y5SuprailiacSkf
Y5AbdominalSkf
Y5BMIz
Y1BMIz
PGS6
PGS7
PGS5
smkPreg
pregWt
Y1And5HeadC
PGS3_PGS4
PGS1_PGS2
Y17 BMI
occupM0
ageMy
htM
wtF
htF
birthYr
birthHeadC
solid
cc
pregWt
Y1SprlSkf
Y1AdbmSkf
Y5MidArmCirc
Y5TricepsSkf
Y5SubscapularSkf
Y5SuprailiacSkf
Y5AbdominalSkf
Y5BMIz
birthBMIz
Y1BMIz
PGS6
PGS7
PGS5
PGS3_PGS4
PGS1_PGS2
Y20 BMI
htM
eduF
wtF
htF
sex
PGS6
PGS5
pregWt
occupM_cc
Y5skf
Y1BMIz
Y5BMIz_Y5ArmChestC
PGS3_PGS4
PGS1_PGS2_PGS7
Y23 BMI
htM
wtF
apgar
placWt
PGS5
pregWt
bf
Y5skf
birthAnthro
Y5BMIz_Y5ArmChestC
occupM_cc
PGS3_PGS4
PGS1_PGS2_PGS6_PGS7

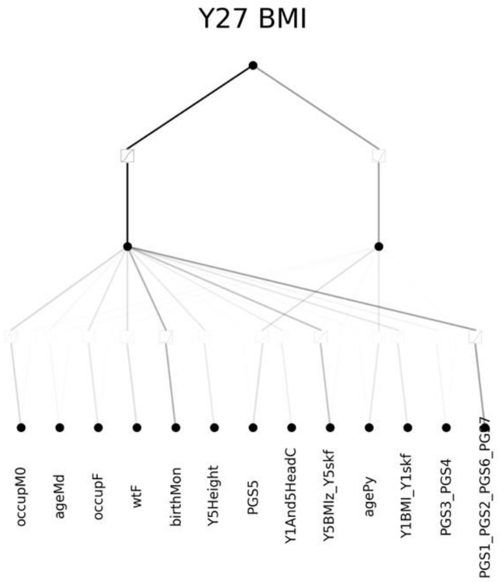


Supplementary Figure 5. Pruned tree plots of the KAN models

### Supplementary 5. BMI estimation and the expression of the key variables

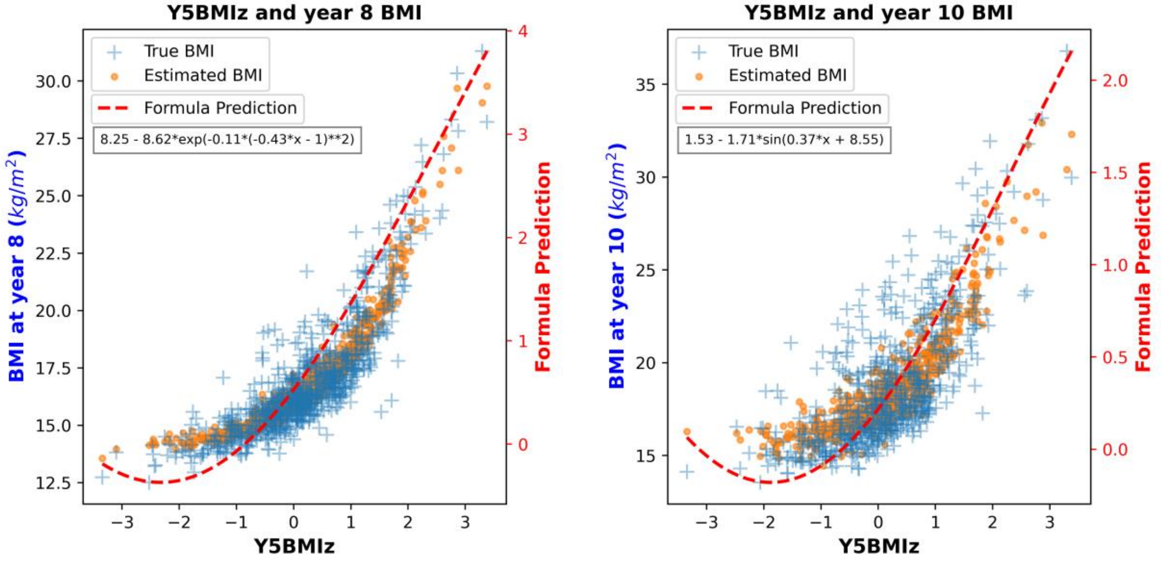

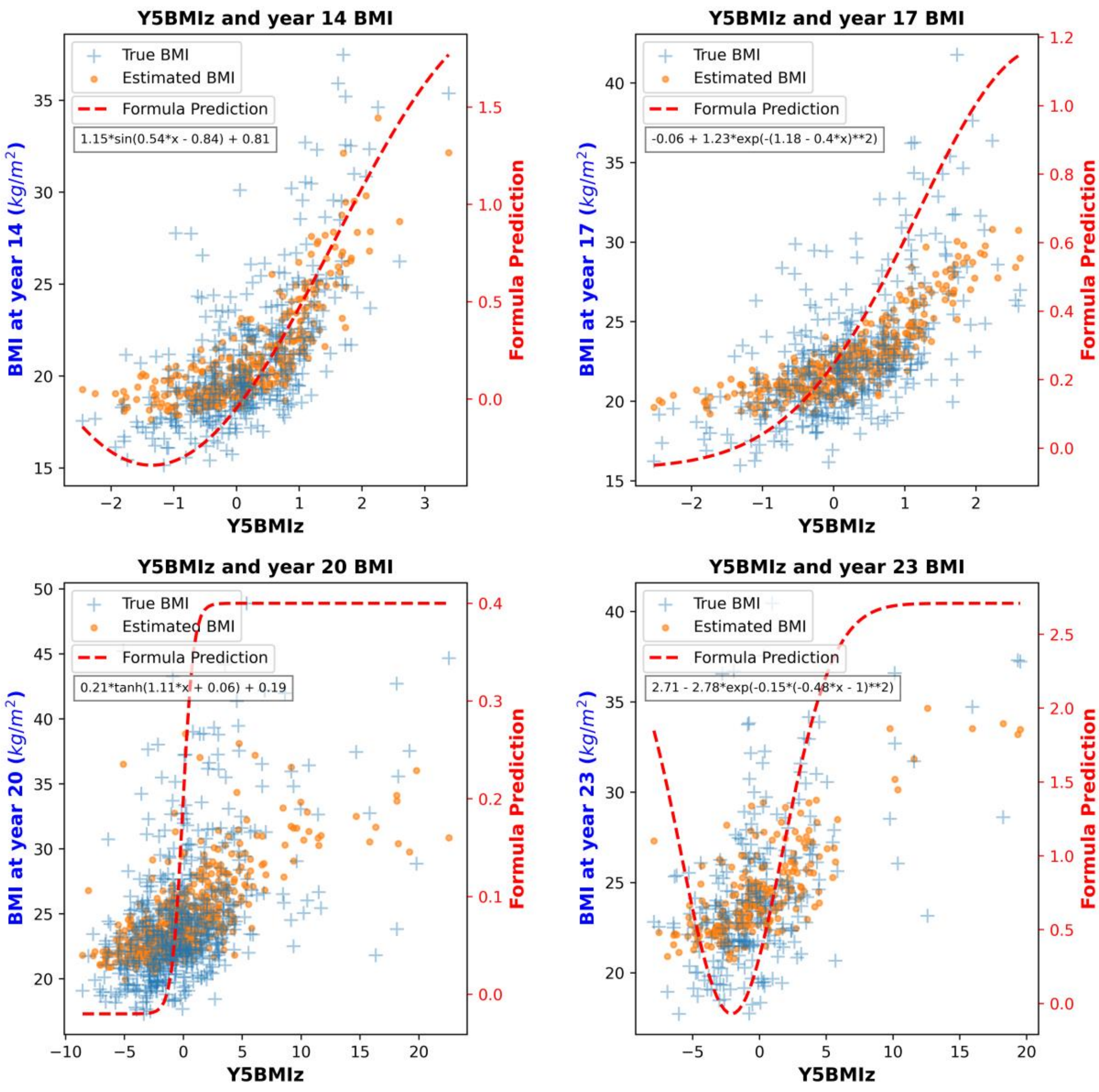


Supplementary Figure 6. BMI estimation and variable formularization on Y5BMIz across the age groups of 8, 10, 14, 17, 20, and 23 years. Orange dots – estimated BMI; blue crosses – real BMI; red dashed lines – formulas of the factor including Y5BMIz; "x" in the legend of function indicates Y5BMIz.

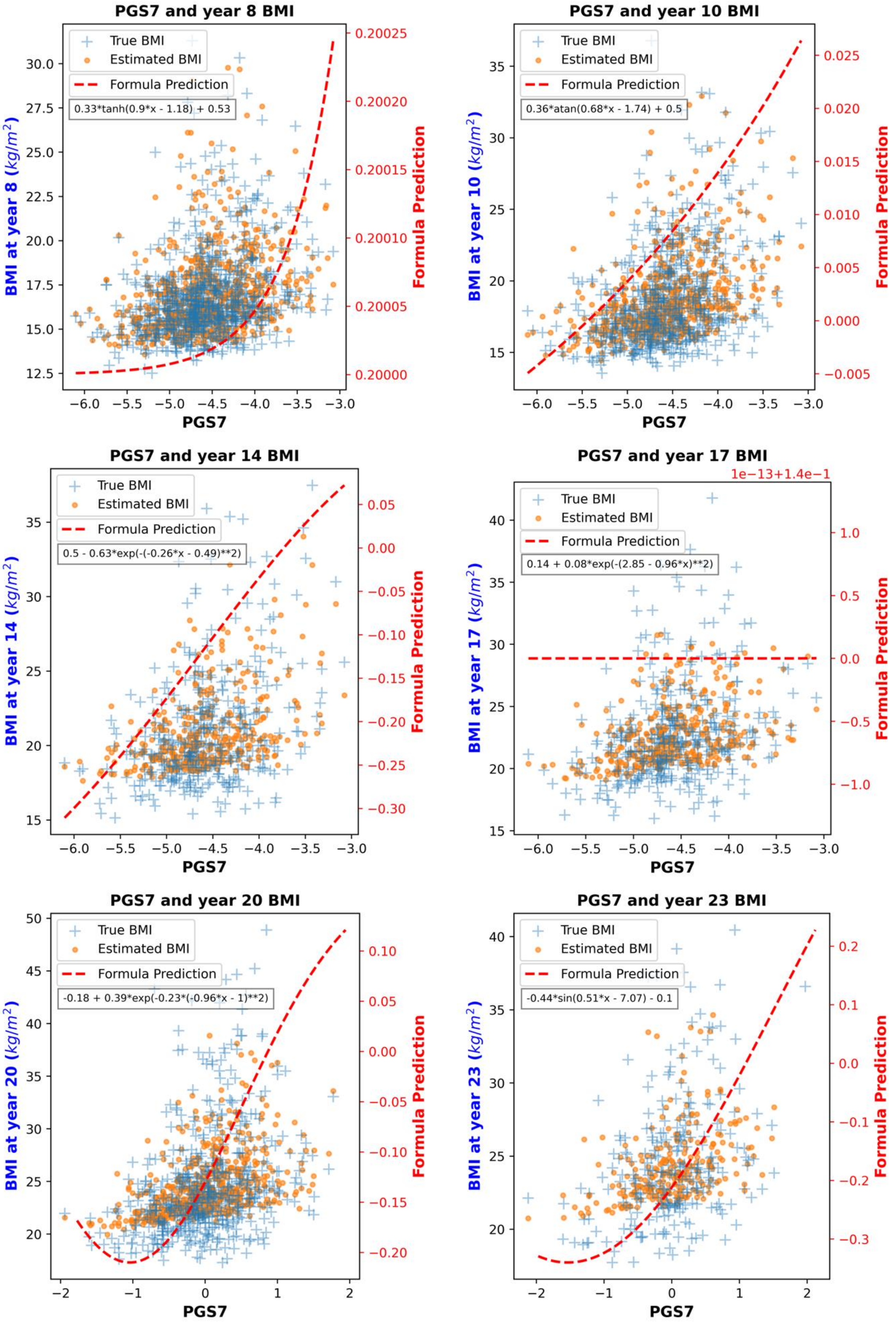
PGS7 and year 8 BMI
True BMI
Estimated BMI
Formula Prediction
0.33*tanh(0.9*x - 1.18) + 0.53
BMI at year 8 (kg/m^2)
Formula Prediction
PGS7
PGS7 and year 10 BMI
True BMI
Estimated BMI
Formula Prediction
0.36*atan(0.68*x - 1.74) + 0.5
BMI at year 10 (kg/m^2)
Formula Prediction
PGS7
PGS7 and year 14 BMI
True BMI
Estimated BMI
Formula Prediction
0.5 - 0.63*exp(-(-0.26*x - 0.49)**2)
BMI at year 14 (kg/m^2)
Formula Prediction
PGS7
PGS7 and year 17 BMI
1e−13+1.4e−1
True BMI
Estimated BMI
Formula Prediction
0.14 + 0.08*exp(-(2.85 - 0.96*x)**2)
BMI at year 17 (kg/m^2)
Formula Prediction
PGS7
PGS7 and year 20 BMI
True BMI
Estimated BMI
Formula Prediction
-0.18 + 0.39*exp(-0.23*(-0.96*x - 1)**2)
BMI at year 20 (kg/m^2)
Formula Prediction
PGS7
PGS7 and year 23 BMI
True BMI
Estimated BMI
Formula Prediction
-0.44*sin(0.51*x - 7.07) - 0.1
BMI at year 23 (kg/m^2)
Formula Prediction
PGS7

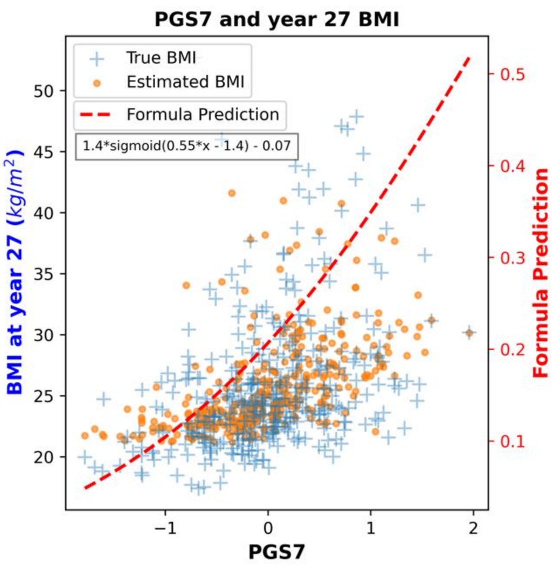


Supplementary Figure 7. BMI estimation and variable formularization on PGS7 across the age groups of 8, 10, 14, 17, 20, 23, and 27 years. Orange dots – estimated BMI; blue crosses – real BMI; red dashed lines – formulas of the factor including P0921; “x” in the legend of function – the factor containing PGS7.
